# Attenuation of antibody titres during 3-6 months after the second dose of the BNT162b2 vaccine depends on sex, with age and smoking as risk factors for lower antibody titres at 6 months

**DOI:** 10.1101/2021.11.14.21266334

**Authors:** Yushi Nomura, Michiru Sawahata, Yosikazu Nakamura, Ryousuke Koike, Otohiro Katsube, Koichi Hagiwara, Seiji Niho, Norihiro Masuda, Takaaki Tanaka, Kumiya Sugiyama

**Affiliations:** Department of Respiratory Medicine and Clinical Immunology, National Hospital Organization Utsunomiya National Hospital; Department of Pulmonary Medicine and Clinical Immunology, Dokkyo Medical University; Division of Pulmonary Medicine, Department of Medicine, Jichi Medical University; Department of Public Health, Jichi Medical University; Department of Surgery, National Hospital Organization Utsunomiya National Hospital; Department of Orthopaedic Surgery, National Hospital Organization Utsunomiya National Hospital; Department of Respiratory Medicine and Clinical Immunology, Dokkyo Medical University Saitama Medical Center

**Keywords:** SARS CoV-2, viral infection, clinical epidemiology

## Abstract

**Objective:** We aimed to determine antibody titres at 6 months and their rate of change during 3-6 months after the second dose of the BNT162b2 coronavirus disease 2019 (COVID-19) mRNA vaccine (Pfizer/BioNTech) and to explore clinical variables associated with titres in Japan.

**Methods:** We enrolled 365 healthcare workers (250 women, 115 men) whose 3-month antibody titres were analyzed in our previous study and whose blood samples were collected 183 ± 15 days after the second dose. Participant characteristics collected previously were used. The relationships of these factors with antibody titres at 6 months and rates of change in antibody titres during 3-6 months were analyzed.

**Results:** Median age was 44 years. Median antibody titre at 6 months was 539 U/mL. Older participants had significantly lower antibody titres (20s, 752 U/mL; 60s–70s, 365 U/mL). In age-adjusted analysis, smoking was the only factor associated with lower antibody titres. Median rate of change in antibody titres during 3-6 months was −29.4%. The only factor significantly associated with the rate of change in Ab titres was not age or smoking, but sex (women, −31.6%; men, −25.1%).

**Conclusion:** The most important factors associated with lower antibody titres at 6 months were age and smoking, as at 3 months, probably reflecting their effect on peak antibody titres. However, antibody titres significantly attenuated during 3-6 months in women alone, which reduced the sex difference in antibody titres seen during the first 3 months. Antibody titres may be affected by different factors at different time points.

## INTRODUCTION

The BNT162b2 vaccine (Pfizer/BioNTech), which was the first coronavirus disease 2019 (COVID-19) mRNA vaccine to be administered to healthcare professionals in Japan, starting in February 2021, is expected to help to prevent both the onset [1] and progression of COVID-19. To obtain the maximum efficacy of vaccination with reduced adverse effects and to optimize and individualize the vaccination regimen, we need to record the exact chronological changes in real-world antibody (Ab) titres following BNT162b2 vaccination and to investigate the relationships of recipients’ medical histories and demographic characteristics with Ab titres.

Factors associated with the rate of change of Ab titres during the first several months after the second of the two BNT162b2 doses could differ from those associated with peak Ab titres immediately after the second of the two BNT162b2 doses. The median titre shortly after the full schedule of this vaccine in Japan has been reported to be 2060 U/mL (interquartile range [IQR], 1250–2650 U/mL) [2], which is similar to the median titre reported in Italy (1975 U/mL; IQR, 895–3455 U/mL) [3]. Various factors, including older age [2, 4-6], male sex [2], ethnicity [7], social condition [7], obesity [8, 9], smoking habit [6, 9], drinking habit [2], hypertension [9], cancer [10, 11], and use of immunosuppressive drugs [2], have been reported to reduce the Ab titres obtained shortly after the second dose of the BNT162b2 vaccine. In Japan, older age, male sex, and drinking habit are reported risk factors for a lower peak value of the Ab titre [2].

In our previous cohort study of the serological Ab titres of 378 healthcare workers at 3 months after the second dose of the BNT162b2 vaccine [12] in Tochigi prefecture, Japan, these titres ranged from 3 to 5790 U/mL, and the median (IQR) Ab titre was 764 (423–1140) U/mL, which was much lower than the above-mentioned value obtained shortly after the second inoculation. We thus determined that age and smoking status are predictors of Ab titres 3 months after the second dose of the BNT162b2 vaccine in Japan. However, the medium-term attenuation rates of Ab titres several months after the second BNT162b2 dose and the relationship of these rates with clinical background and demographic characteristics are still not known.

Against this background, we analyzed Ab titres at 6 months and their rate of change during 3-6 months after the second dose of the BNT162b2 vaccine and explored the clinical variables associated with these parameters in Japan.

## METHODS

### Population and study design

In this single-center prospective observational study, we recruited healthcare workers whose blood samples were collected 183 ± 15 days after the second of two BNT162b2 vaccine inoculations (Pfizer/BioNTech) administered 3 weeks apart in February and March 2021 in the National Hospital Organization Utsunomiya National Hospital in Tochigi prefecture, Japan.

Initially, we recruited 378 participants whose Ab titres 3 months after the second dose of the BNT162b2 COVID-19 vaccine were already analyzed in our previous study [12]. Their medical histories and demographic characteristics were recorded in that study using a structured self-report questionnaire [12]. However, we excluded 10 participants whose blood samples were not obtained at 6 months and 3 participants whose blood sampling confirmed the presence of Abs against the nucleocapsid proteins for severe acute respiratory syndrome coronavirus 2 (SARS-CoV-2). Finally, we enrolled 365 healthcare workers (250 women, 115 men).

Blood samples collected 183 ± 15 days after the second inoculation were used to measure total Ab titres against the SARS-CoV-2 spike antigen using a commercially available electrochemiluminescence immunoassay (ECLIA) (Elecsys® Anti-SARS-CoV-2 RUO; Roche Diagnostics) [13]. The relationships between Ab titres against the SARS-CoV-2 spike antigen and clinical and lifestyle characteristics were analyzed. In age-adjusted analysis, individual Ab titres were recalculated by subtracting the median Ab titre of the corresponding age group from an individual’s Ab titre. For example, an age-adjusted Ab titre for an individual in his/her 20s was calculated as follows: individual Ab titre -median Ab titre for participants in their 20s.

In addition, we calculated the rate of change in Ab titres during 3-6 months after the second dose of the BNT162b2 COVID-19 vaccine. Individual rates of change in Ab titres were calculated as follows: rate of change = [(Ab titre 6 months after the 2nd dose - Ab titre 3 months after the 2nd dose [12]) / Ab titre 3 months after the 2nd dose] × 100 (%). Then, the relationships between these rates of change in Ab titres against the SARS-CoV-2 spike antigen and clinical and lifestyle characteristics were analyzed.

The Ethics Committee of National Hospital Organization Utsunomiya National Hospital (No. 03-01; April 19, 2021) approved this study, and written informed consent was obtained from all participants before enrollment.

### Data analysis

Nonparametric continuous data are expressed as the median with the IQR. Categorical data are presented as absolute numbers and relative frequencies (*n*, %). To calculate Spearman’s rank correlation coefficient and perform the Mann–Whitney *U* test, we used Statistical Package for the Social Sciences (SPSS version 25).

## RESULTS

### Study population

In total, 365 healthcare workers (250 women, 115 men) were enrolled in this study. Their baseline characteristics are summarized in **Table 1**. Briefly, the median (IQR) age of the participants was 44 (32–54) years. Nurses (n=171) and physicians (n=34) comprised 56.2% of the study population.

**Table 1.** Participants’ baseline characteristics (*N*=365)

| Variable | Total | Ab titre, median (IQR), U/mL | Correlation coefficient $\rho$ | P-value |
| --- | --- | --- | --- | --- |
| Age, median (IQR), y | 44 (32-54) |  | -0.347 | <0.0001<sup>#</sup> |
| Sex (M/F), *n* | 115/250 | 482 (305 to 865) / 572 (316 to 884) |  | 0.1970<sup>*</sup> |
| Body mass index, median (IQR), kg/m<sup>2</sup> | 22.4 (20.2-24.8) |  | -0.017 | 0.7446<sup>#</sup> |
| Smoking (ever/never), *n* | 149/216 | 406 (213 to 688) / 614 (405 to 957) |  | <0.0001<sup>*</sup> |
| current smoker/ ex smoker/ unknown | 90/45/14 | 354 (174 to 564) / 436 (225 to 894)<sup>§§</sup> |  | 0.0609<sup>*</sup> |
|  |  | ex smoker vs never smoker |  | 0.0014<sup>*</sup> |
|  |  | current smoker vs never smoker |  | <0.0001<sup>*</sup> |
| Brinkman Index<sup>§§, §§§</sup>, *n* | 129 |  | -0.197 | 0.0258<sup>#</sup> |
| Drinking, *n* | 228/134/3<sup>§</sup> | 544 (299 to 881)/ 536 (348 to 868)<sup>§§</sup> |  | 0.7697<sup>*</sup> |
| Allergy, *n* |  |  |  |  |
| Food | 38/292/35<sup>§</sup> | 515 (307 to 925) / 544 (318 to 807)<sup>§§</sup> |  | 0.7697<sup>*</sup> |
| Drug | 37/293/35<sup>§</sup> | 517 (287 to 759) / 549 (321 to 833)<sup>§§</sup> |  | 0.5858<sup>*</sup> |
| Allergic disease, *n* |  |  |  |  |
| Allergic rhinitis including pollinosis | 167/175/23<sup>§</sup> | 546 (258 to 895) / 523 (320 to 792)<sup>§§</sup> |  | 0.8242<sup>*</sup> |
| Bronchial asthma | 43/299/23<sup>§</sup> | 582 (360 to 885) / 522 304 to 837)<sup>§§</sup> |  | 0.5643<sup>*</sup> |
| Skin allergy including atopic dermatitis | 47/295/23<sup>§</sup> | 641(380 to 1005) / 522(303 to 815)<sup>§§</sup> |  | 0.0710<sup>*</sup> |
| Diabetes mellitus, *n* | 12/340/13<sup>§</sup> | 331 (149 to 589) / 540 (318 to 878)<sup>§§</sup> |  | 0.0512<sup>*</sup> |
| Hypertension, *n* | 27/325/13<sup>§</sup> | 356 (195 to 566) / 553 (321 to 872)<sup>§§</sup> |  | 0.0327<sup>*</sup> |
| Dyslipidaemia, *n* | 18/334/13<sup>§</sup> | 351 (205 to 698) / 544 (317 to 877)<sup>§§</sup> |  | 0.0834<sup>*</sup> |
| Collagen disease, *n* | 13/332/20<sup>§</sup> | 303 (126 to 840) / 544 (324 to 843)<sup>§§</sup> |  | 0.2695<sup>*</sup> |

IQR=interquartile range
<sup>\*</sup>: Mann-Whitney *U* test
<sup>#</sup>: Spearman's rank correlation coefficient test
<sup>§</sup>: yes/no/unknown are shown
<sup>§§</sup>: unknown is excluded
<sup>§§§</sup>: ever smoker only

### Distribution of Ab titres against the SARS-CoV-2 spike antigen 6 months after the second dose of the BNT162b2 COVID-19 vaccine by age and sex

The median (IQR) Ab titre against the SARS-CoV-2 spike antigen 6 months after vaccination with the BNT162b2 COVID-19 vaccine was 539 (309-872) U/mL. Older participants had significantly lower SARS-CoV-2 Ab titres (correlation coefficient ρ = −0.347) (**Table 1** and **Figure 1**). Ab titres tended to decrease in both men and women as participants’ age increased from their 20s to 70s. Median antibody titres of men in their 20s, 30s, 40s, 50s, and 60s–70s were 823, 648, 485, 335, and 432 U/mL, respectively, whereas median Ab titres of women in their 20s, 30s, 40s, 50s, and 60s–70s were 748, 641, 560, 490, and 318 U/mL, respectively.

**Figure 1.**
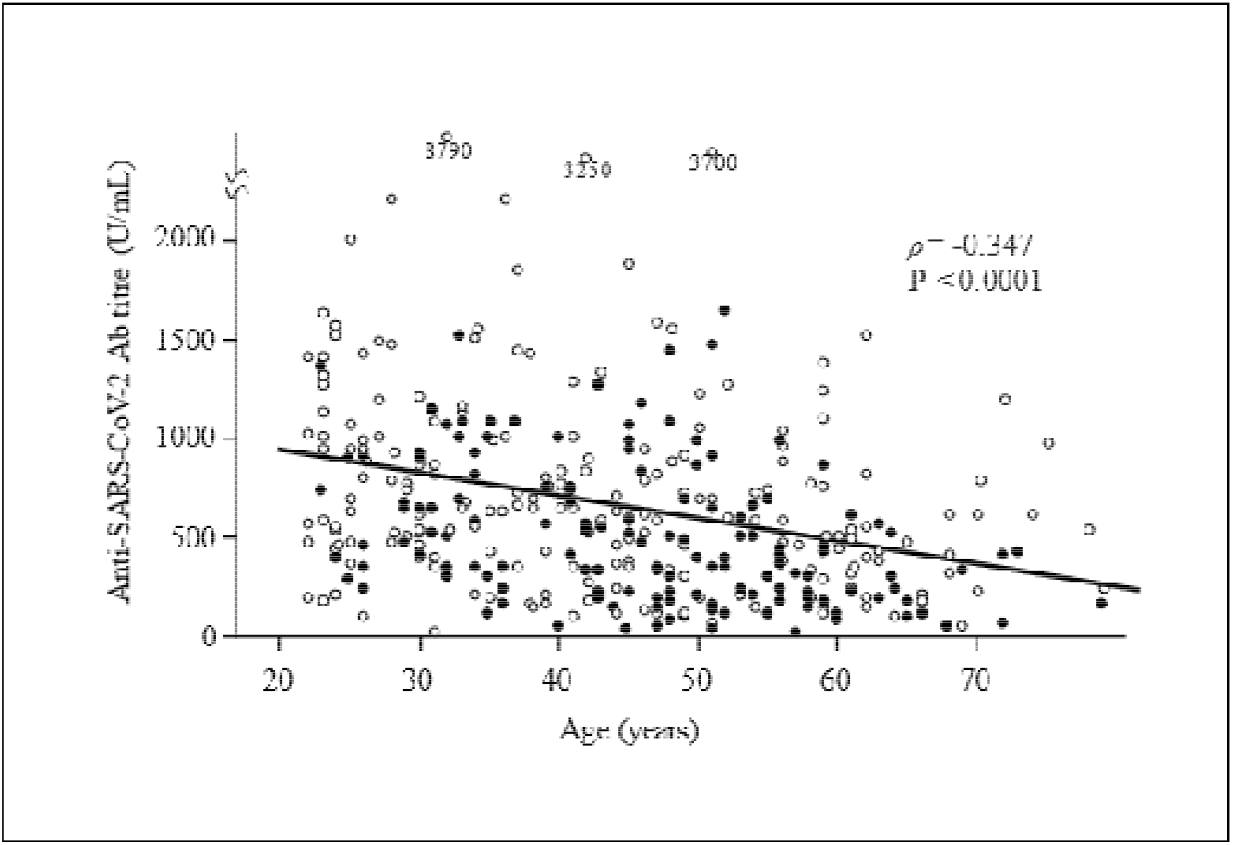
Scatter plot of the distribution of Ab titres 6 months after the second vaccine dose according to age and smoking status. Older participants had significantly lower Ab titres. Closed and open circles show ever-smokers and never-smokers, respectively. Ever-smokers had lower Ab titres than never-smokers.

### Relationship between the Ab titres against the SARS-CoV-2 spike antigen 6 months after vaccination and risk factors

We first performed univariate analyses to identify factors associated with serum Ab titres against the SARS-CoV-2 spike protein. The factors significantly associated with lower Ab titres were older age, smoking, and hypertension (**Table 1**). Ever-smokers tended to have lower Ab titres than never-smokers (**Figure 1**).

We also analyzed the risk factors for lower Ab titres after adjustment for age because the prevalence of certain factors may differ according to age, such as hypertension. In the age-adjusted analysis, individual Ab titres were recalculated by subtracting the median Ab titre of the corresponding age group from an individual’s Ab titre. Median Ab titres of participants in their 20s, 30s, 40s, 50s, and 60s–70s were 752, 642, 550, 418, and 365 U/mL, respectively. Thus, the age-adjusted Ab titre for an individual in his/her 20s was calculated as “individual Ab titre - 752”. After age adjustment, the only factor significantly associated with lower Ab titres was smoking (**Table 2**). In terms of smoking, the age-adjusted median (IQR) Ab titres were −97 (−277 to 184) and 56 (−182 to 342) in ever-smokers and never-smokers, respectively.

**Table 2.** Age-adjusted data of median Ab titres (*N*=365)

| Variable | Ab titre, median (IQR), U/mL | Correlation coefficient $\rho$ | P-value |
| --- | --- | --- | --- |
| Male/female | -15 (-246 to 241) / 8 (-209 to 290) |  | 0.2851 * |
| Body mass index, median (IQR), kg/m<sup>2</sup> |  | 0.023 | 0.6647 # |
| Smoking (ever/never) | -97 (-277 to 184) / 56 (-182 to 342) |  | <0.0001 * |
| current smoker/ ex smoker | -205 (-320 to 7) / -72 (-264 to 256) |  | 0.0255 * |
| Brinkman Index §§, §§§ | ex smoker vs never smoker<br>current smoker vs never smoker | 0.012 | 0.0203 *<br><0.0001 *<br>0.8935 # |
| Drinking | 3 (-242 to 296) / 5 (-205 to 249) §, §§ |  | 0.6475 * |
| Allergy |  |  |  |
| Food | 18 (-250 to 235) / -2 (-223 to 256) §, §§ |  | 0.9604 * |
| Drug | -4 (-131 to 271) / -1 (-238 to 251) §, §§ |  | 0.5858 * |
| Allergic disease |  |  |  |
| Allergic rhinitis including pollinosis | -7 (-240 to 271) / -1 (-223 to 253) §, §§ |  | 0.8473 * |
| Bronchial asthma | 30 (-223 to 254) / -8 (-239 to 263) §, §§ |  | 0.7390 * |
| Skin allergy including atopic dermatitis | 61 (-166 to 421) / -9 (-240 to 248) §, §§ |  | 0.1638 * |
| Diabetes mellitus | -60 (-269 to 77) / 0 (-225 to 272) §, §§ |  | 0.4696 * |
| Hypertension | -69 (-247 to 183) / 0 (-225 to 271) §, §§ |  | 0.6409 * |
| Dyslipidaemia | -56 (-220 to 259) / 0 (-232 to 268) §, §§ |  | 0.7392 * |
| Collagen disease | -227 (-368 to 290) / 0 (-222 to 261) §, §§ |  | 0.3021 * |

IQR=interquartile range
\*: Mann–Whitney *U* test

### Distribution of the rate of change in Ab titres during 3-6 months after the second dose of the BNT162b2 COVID-19 vaccine by age and sex

The median (IQR) rate of change in Ab titres against the SARS-CoV-2 spike antigen during 3-6 months after vaccination was −29.4% (−40.4% to −17.7%). No significant correlation was observed between the rate of change in the anti-SARS-CoV-2 Ab titre and age (correlation coefficient ρ = −0.028) (**Table 3** and **Figure 2A**). Median rates of change in Ab titres of men in their 20s, 30s, 40s, 50s, and 60s–70s were −24.3, −25.3, −24.0, −33.7, and −14.8 U/mL, respectively, whereas those of women in their 20s, 30s, 40s, 50s, and 60s–70s were −33.8, −31.5, −31.5, −30.0, and −32.6 U/mL, respectively. Surprisingly, the only factor significantly associated with the rate of change in the Ab titre was not age or smoking, but sex (**Table 3**). In terms of sex, median (IQR) rates of change in Ab titres were −31.6% (−42.0% to −20.2%) and −25.1% (−36.8% to −12.0%) in women and men, respectively (**Table 3**), suggesting that Ab titres decrease faster in women than in men.

**Table 3.** Median rate of change in Ab titres from 3 months to 6 months (*N*=365)

| Variable | Ab titre, median (IQR), % <sup>§</sup> | Correlation coefficient $\rho$ | P-value |
| --- | --- | --- | --- |
| Age |  | -0.028 | 0.5876 # |
| Male/female | -25.1 (-36.8 to -12.0) / -31.6 (-42.0 to -20.2) |  | 0.0005 * |
| Body mass index, median (IQR), kg/m<sup>2</sup> |  | 0.025 | 0.6274 # |
| Smoking (ever/never)<br>current smoker/ ex smoker | -28.4 (-39.7 to -15.5) / -30.3 (-40.7 to -19.0)<br>-31.7 (-40.6 to -18.3) / -27.4 (-40.1 to -16.1) <sup>§§</sup><br>ex smoker vs never smoker |  | 0.3051 *<br>0.3853 *<br>0.2914 * |
| Brinkman Index <sup>§§, §§§</sup> | current smoker vs never smoker | -0.034 | 0.8809 *<br>0.6990 # |
| Drinking | -29.1(-41.0 to -17.8) / -30.5(-39.1 to -18.1) <sup>§, §§</sup> |  | 0.777 * |
| Allergy |  |  |  |
| Food | -29.4 (-37.1 to -16.5) / -29.4 (-39.9 to -18.6) <sup>§, §§</sup> |  | 0.7821 * |
| Drug | -28.8 (-40.7 to -22.6) / -29.7 (-39.3 to -18.3) <sup>§, §§</sup> |  | 0.7469 * |
| Allergic disease |  |  |  |
| Allergic rhinitis including pollinosis | -29.2 (-40.9 to -18.1) / -29.6 (-39.6 to -18.0) <sup>§, §§</sup> |  | 0.8606 * |
| Bronchial asthma | -31.7 (-40.5 to -19.4) / -29.1 (-40.5 to -17.6) <sup>§, §§</sup> |  | 0.5521 * |
| Skin allergy including atopic dermatitis | -30.3 (-44.6 to -25.1) / -29.1 (-39.7 to -17.6) <sup>§, §§</sup> |  | 0.1677 * |
| Diabetes mellitus | -27.6 (-32.8 to -11.4) / -29.4 (-40.5 to -18.0) <sup>§, §§</sup> |  | 0.3617 * |
| Hypertension | -24.0 (-32.1 to -12.3) / -29.7 (-40.8 to -18.0) <sup>§, §§</sup> |  | 0.0679 * |
| Dyslipidaemia | -31.7 (-39.2 to -22.8) / -29.3 (-40.5 to -17.7) <sup>§, §§</sup> |  | 0.7178 * |
| Collagen disease | -27.0 (-48.6 to -19.1) / -29.6 (-40.3 to -17.9) <sup>§, §§</sup> |  | 0.8895 * |

IQR=interquartile range
\*: Mann–Whitney *U* test

**Figure 2.**
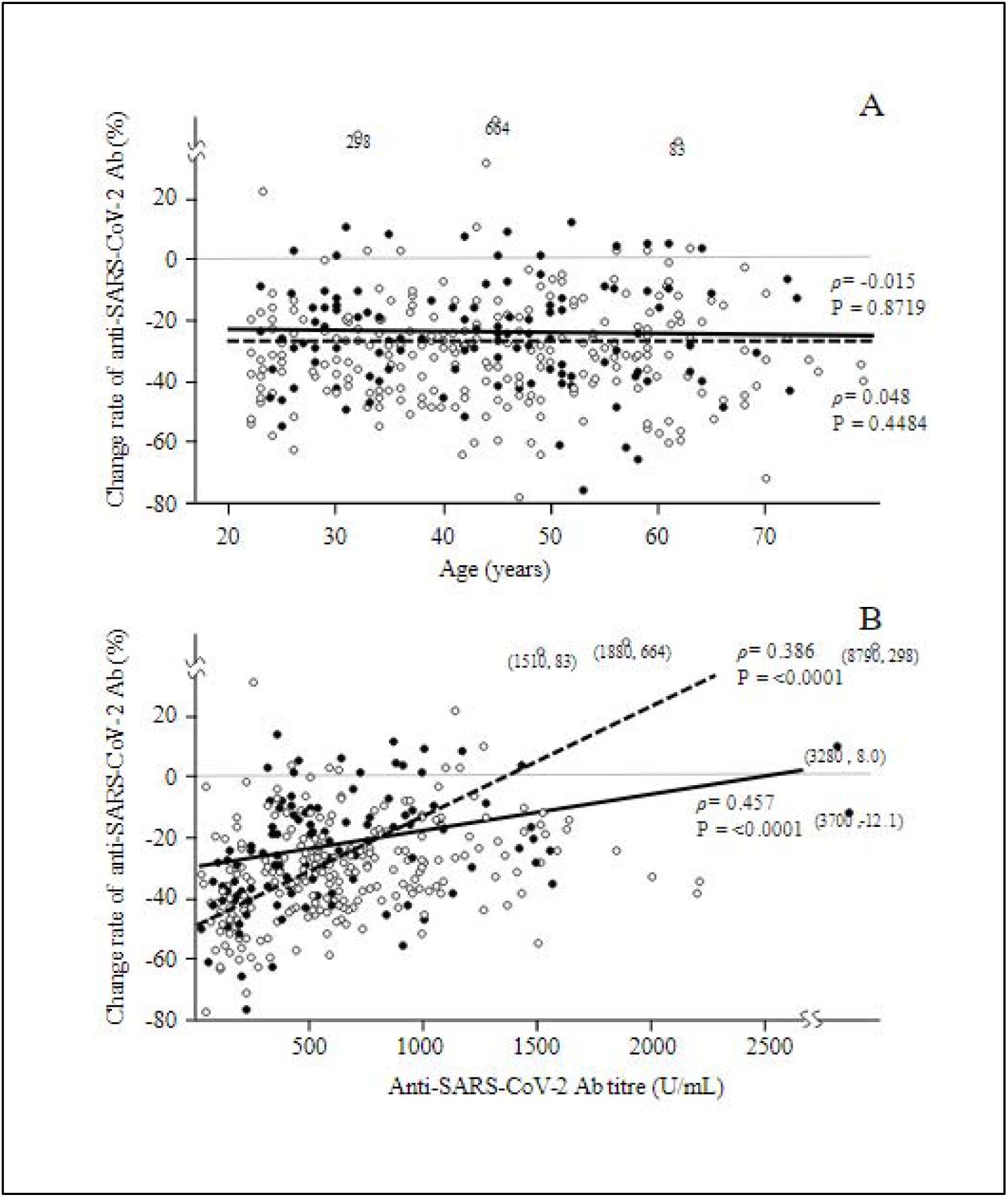
Scatter plot of the distribution of the rate of change in Ab titres during 3-6 months after the second dose of the vaccine according to sex. The relationship between the rate of change after the vaccination and age is shown in A, and the relationship between the rate of change and the Ab titre 6 months after the vaccination is shown in B. No significant correlation was observed in A, and age did not affect the attenuation of the Ab titres from 3 to 6 months after the vaccination. However, a significant correlation was observed in B, and a lower Ab titre might affect the attenuation of Ab titres from 3 to 6 months after the vaccination. Closed and open circles and continuous and broken lines show men and women, respectively.

An additional analysis was performed (**Table 4**) because smoking was an important risk factor at both 3 and 6 months after vaccination and the smoking rates differed between men and women. For age-adjusted median Ab titres, no significant sex differences were observed in the ever-smoker and never-smoker groups. However, both the male and female groups showed significant differences by smoking status in age-adjusted median Ab titres. On the other hand, no significant differences in the median rate of change in Ab titres by smoking status were observed in the male and female groups. However, both the ever-smoker and never-smoker groups showed significant sex differences in the median rate of change in Ab titres.

**Table 4.**
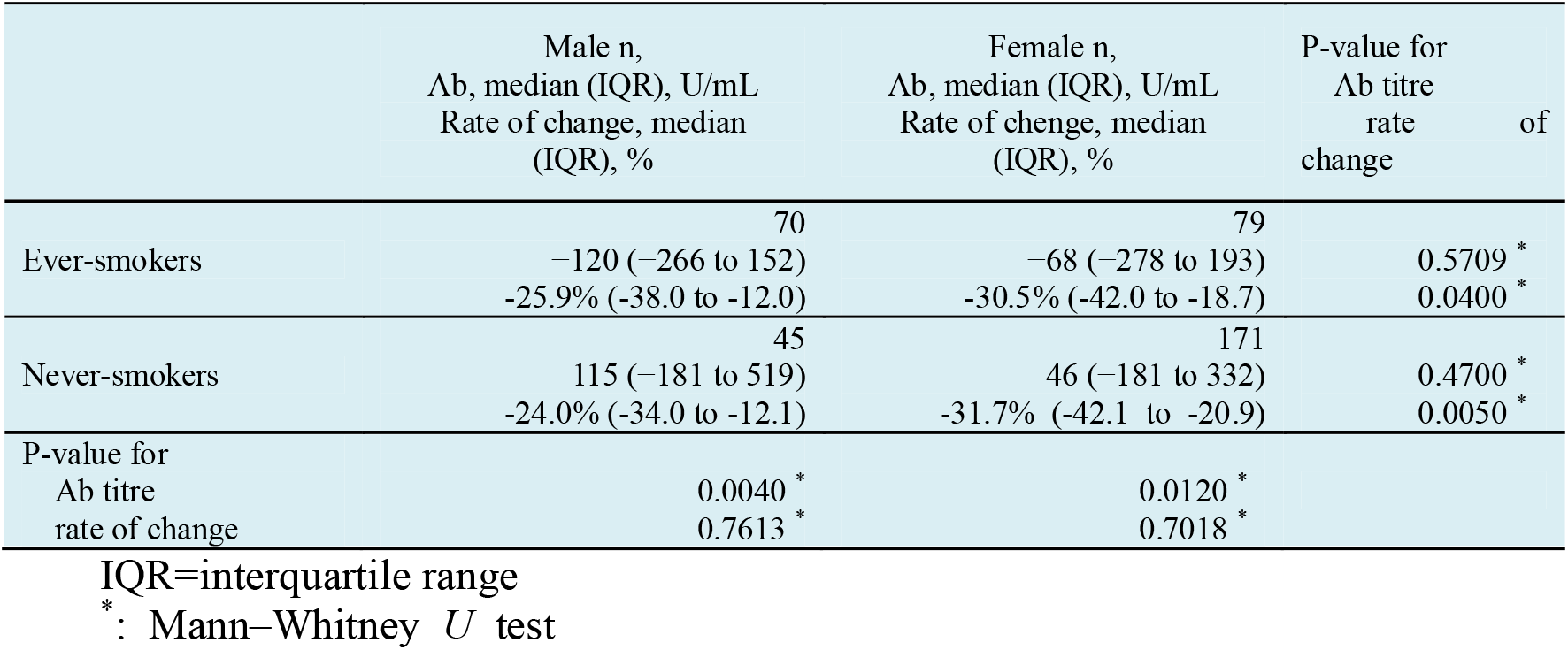
Age-adjusted median Ab titres and rates of change in the relationship between sex and smoking.

|  | Male n,<br>Ab, median (IQR), U/mL<br>Rate of change, median<br>(IQR), % | Female n,<br>Ab, median (IQR), U/mL<br>Rate of change, median<br>(IQR), % | P-value for<br>Ab titre<br>rate of<br>change |
| --- | --- | --- | --- |
| Ever-smokers | 70<br>-120 (-266 to 152)<br>-25.9% (-38.0 to -12.0) | 79<br>-68 (-278 to 193)<br>-30.5% (-42.0 to -18.7) | 0.5709 *<br>0.0400 * |
| Never-smokers | 45<br>115 (-181 to 519)<br>-24.0% (-34.0 to -12.1) | 171<br>46 (-181 to 332)<br>-31.7% (-42.1 to -20.9) | 0.4700 *<br>0.0050 * |
| P-value for<br>Ab titre<br>rate of change | 0.0040 *<br>0.7613 * | 0.0120 *<br>0.7018 * |  |

IQR=interquartile range \*: Mann–Whitney *U* test

To define the factors affecting the rates of change in Ab titres, the relationship between the rates of change in Ab titres and the Ab titre 6 months after vaccination was analyzed (**Figure 2B**) because the Ab titres 3–6 months after vaccination were significantly higher in women than in men. A significant positive correlation was observed, indicating that participants with a lower Ab titre had greater attenuation of the Ab titre.

## DISCUSSION

To our knowledge, this is the first study to report real-world Ab titres against the SARS-CoV-2 spike antigen at 6 months and the rates of change in Ab titres during 3-6 months after vaccination and to identify the factors associated with these parameters from a comprehensive range of clinical and lifestyle characteristics in Japan. Four important findings were obtained. First, in a cohort with a median (IQR) age of 44 (32–54) years, the median Ab titre 6 months after the second dose was 539 U/mL. Older participants had significantly lower Ab titres, with an Ab titre about half that of those in their 20s. Second, in an age-adjusted analysis, the only risk factor for a lower Ab titre was smoking. Third, the median rate of change in the Ab titre during 3-6 months after vaccination was −29.4%. Surprisingly, the only factor significantly associated with attenuation of the Ab titres was not age or smoking, but sex. The median rates of change in Ab titres were −31.6% and −25.1% in women and men, respectively. Fourth, both male and female participants with a lower Ab titre showed greater attenuation of the Ab titre. This means that the rate of change in Ab titres may be affected by individual differences.

In terms of the first and second findings, the most important factors associated with a low Ab titre 6 months after the second dose were still age and smoking habit, as seen 3 months after the second dose, probably reflecting their effect on peak Ab titres. Interestingly, a reversal of the sex difference in Ab titres seen 3 months after the second dose was observed at 6 months; in our previous study, the median Ab titre was 764 U/mL 3 months after the second dose [12]. The median Ab titres were 942 and 1095 U/mL in men and women in their 20s, respectively, but 490 and 519 U/mL in men and women in their 60s–70s, respectively. In the age-adjusted analysis, the only risk factors for a lower Ab titre 3 months after the second dose [12] were male sex and smoking. However, we concluded that the sex difference may have arisen from the sex difference in the smoking rate, rather than from biological sex differences.

Regarding the third finding, the only factor significantly associated with the attenuation in Ab titres during 3-6 months after the second dose was not age or smoking but sex, which seemingly resulted in a reduction of the sex difference in Ab titres seen 3 months after the second dose. This suggests that Ab titres decrease faster in women than in men.

Concerning the fourth finding, we hypothesized that participants with a higher Ab titre would show greater attenuation of Ab titres because the rate of change during 3-6 months and the Ab titre 3 months after vaccination were significantly higher in women than in men. However, a significant positive correlation was observed between the rate of change in Ab titres and the Ab titre 6 months after vaccination. The Ab titre at 6 months after vaccination was associated with the rate of attenuation during 3-6 months, and participants with a lower Ab titre showed greater attenuation of the titre. We could not find other risk factors, such as age and comorbidities, beyond the above. Although we concluded that the attenuation was affected by individual differences, we will continue to define the risk factors through additional study.

The factors identified to affect Ab titres vary by the time elapsed since vaccination. For peak Ab titre 2 to 5 weeks after vaccination, the reported risk factors were sex, age, and alcohol intake. Three months after vaccination, age and smoking were risk factors in our study, and the effects of sex and alcohol intake were not observed. Six months after vaccination, age and smoking were still risk factors in our study. However, these factors did not affect the attenuation during 3-6 months. This means that these factors will strongly affect the Ab titre within 3 months after vaccination and that the effects will persist 6 months after vaccination. From 3 to 6 months after vaccination, sex and lower Ab titre were risk factors. According to the above results, smoking is the most important factor that could be avoided to maintain a higher Ab titre. Thus, it appears that different factors affect the Ab titre at different time points.

Some limitations and possible sources of bias in this study include the following. First, the participants were limited in number and were all healthcare workers vaccinated at a single national hospital in Tochigi prefecture. Therefore, the results obtained in this study might not be generalizable on a wide scale, or even within Japan. Second, we excluded participants with Abs against nucleocapsid proteins on the presumption that they had previously been infected with COVID-19. However, some of these patients had slight increases in Ab titres against the spike protein but became negative for Ab titres against nucleocapsid proteins. One possibility is that they had not been infected with COVID-19 and that the Ab titres against the spike protein had been increased by individual differences. The other possibility is that the Ab titres against nucleocapsid proteins might have not increased due to a small viral load of SARS-CoV-2, even though they may still have been infected with COVID-19. To determine the chance of exposure to a small amount of SARS-CoV-2 virus, we analyzed Ab titres in participants who worked in the COVID-19 ward (data not shown). However, their Ab titres were not increased. If the excluded participants were by chance exposed to a small amount of SARS-CoV-2 virus, it may have been through daily life and not the COVID-19 ward. In addition, 5 participants had Ab titres exceeding 3000 U/mL and/or a greater than 80% rate of increase in Ab titres against the spike protein, despite negative Abs against nucleocapsid proteins. They may have been infected with COVID-19 considering that a participant with an Ab titre against spike proteins exceeding 5000 U/mL and without Ab titres against nucleocapsid proteins 3 months after vaccination had positive Ab titres against nucleocapsid proteins at 6 months. The participant may have been infected with COVID-19 3 months after vaccination and the Abs against nucleocapsid proteins may have increased at a late stage due to the infection. However, we do not have evidence that the above 5 participants were infected with COVID-19 and we did not exclude them from our analysis.

In conclusion, the most important factors associated with a low Ab titre 6 months after the second dose were age and smoking habit, as seen 3 months after the second dose, probably reflecting the effects of these factors on peak Ab titres. However, the attenuation of Ab titres during 3-6 months after the second BNT162b2 dose depended not on age or smoking, but on sex. A significant attenuation of Ab titres from 3 to 6 months was solely observed in women and resulted in a reduction of the sex difference in the Ab titre seen at 3 months. Further studies of the associations between Ab titres and the comprehensive medical histories of individuals are needed to establish a more personalized approach to vaccination involving earlier boosters, different schedules, or different types of vaccines.

## Data Availability

All data produced in the present work are contained in the manuscript

## Author Contributions

Conceptualisation, M.S., K.S., Y.N. (Yushi Nomura) and Y.N. (Yosikazu Nakamura); methodology and software, Y.N. (Yushi Nomura), K.S., M.S. and Y.N. (Yosikazu Nakamura); validation, K.S., M.S. and Y.N. (Yosikazu Nakamura); formal analysis and investigation, K.S., Y.N. (Yushi Nomura), M.S. and Y.N. (Yosikazu Nakamura); resources, T.T., K.S., N.M., Y.N. (Yushi Nomura), M.S., O.K., and R.K.; data curation, Y.N. (Yushi Nomura), K.S. and M.S.; writing—original draft preparation, M.S., K.S., Y.N. (Yushi Nomura) and Y.N. (Yosikazu Nakamura); writing—review and editing, T.T., N.M., S.N., K.H., O.K., and R.K.; visualisation, K.S., Y.N. (Yushi Nomura) and M.S.; supervision, M.S., K.S., T.T., S.N. and K.H.; project administration, K.S. and T.T.; funding acquisition, T.T. and K.S. All authors have read and agreed to the published version of the manuscript.

## Funding

This research received no external funding.

### Institutional Review Board Statement

The study was conducted according to the guidelines of the Declaration of Helsinki and approved by the Ethics Committee of National Hospital Organization Utsunomiya National Hospital (No. 03-01, 19 April 2021).

### Informed Consent Statement

Informed consent was obtained from all subjects involved in the study.

## Acknowledgments

The authors thank all staff at National Hospital Organization Utsunomiya National Hospital for providing support during sample collection. We would like to thank Miwa Okada, Minako Yamagishi, Junko Shibayama, Yuko Tajima, Mami Ochiai, Midori Takahashi, Hiroko Ueno, Natsuka Suzuki and Yoshino Iwaya for providing support during data analysis. Conflicts of Interest: The authors declare no conflict of interest.

## Abbreviations

COVID-19: coronavirus disease 2019
Ab: antibody
IQR: interquartile range
SARS-CoV-2: severe acute respiratory syndrome coronavirus 2

